# Identifying clinician perceived priorities for a real-time wearable system for in-hospital monitoring: findings and evolutions following the COVID-19 pandemic

**DOI:** 10.64898/2026.04.21.26350610

**Authors:** Sarah Vollam, Cristian Roman, Elizabeth King, Lionel Tarassenko

**Affiliations:** Nuffield Department of Clinical Neurosciences, University of Oxford, Oxford, UK; NIHR Oxford Biomedical Research Centre, Oxford, UK; Institute of Biomedical Engineering, Department of Engineering Science, University of Oxford, Oxford, UK; Oxford Critical Care, Oxford University Hospitals NHS Foundation Trust, Oxford, UK

## Abstract

A Wearable Monitoring System (WMS), comprising a chest patch, wrist-worn pulse oximeter, and arm-worn blood pressure device, was developed in preparation for a pilot Randomised Controlled Trial (RCT) on a UK surgical ward. The system was designed to support continuous physiological monitoring and early detection of deterioration.

An initial prototype user interface was developed by the research team based on prior clinical experience and engineering knowledge. To ensure suitability for clinical practice, iterative user-centred refinement was undertaken through a series of clinician focus groups and wearability assessments. Six focus groups were conducted between November 2019 and May 2021 involving multidisciplinary healthcare professionals. Feedback from these sessions informed successive interface and system modifications.

System development spanned the COVID-19 pandemic, during which the WMS was rapidly adapted and deployed to support clinical care on isolation wards. Feedback obtained during this period was incorporated into later versions of the system and provided a unique opportunity to examine changes in clinician priorities under pandemic conditions.

Clinicians consistently prioritised alert visibility, alarm fatigue mitigation, parameter flexibility, and centralised monitoring. Notably, preferences regarding alert modality and access mechanisms evolved over time: early enthusiasm for mobile or smartphone-type devices shifted towards a preference for fixed, ward-based displays and audible alerts at the nurses’ station following pandemic deployment.

Building on previous wearability testing in healthy volunteers, wearability testing using a validated questionnaire was completed by 169 patient participants during the RCT. The chest patch and pulse oximeter demonstrated high tolerability, whereas the blood pressure cuff showed poor wearability and was removed from the final system.

These findings demonstrate the importance of iterative, clinician-led design for wearable WMS and highlight how extreme clinical contexts such as the COVID-19 pandemic can significantly reshape perceived requirements for safety-critical monitoring technologies.

## 1. INTRODUCTION

Continuous monitoring of hospitalised patients using wearable technologies has the potential to improve early detection of physiological deterioration, reduce unplanned intensive care admissions, and optimise clinical workflows^1^. Advances in wearable sensors, wireless communication, and data visualisation have enabled the development of Wearable Monitoring Systems (WMS) capable of capturing high-resolution physiological data outside traditional high-dependency settings^2,3^. Despite technical feasibility, successful clinical adoption of WMS remains challenging. Systems must balance accuracy, reliability, and data richness with usability, wearability, alarm burden, and integration into existing clinical workflows^1,2,4^. Poorly designed interfaces or alerting strategies can contribute to alarm fatigue, workflow disruption, and reduced clinician trust, ultimately undermining patient safety^5–7^. User-centred design approaches, incorporating clinician feedback throughout development, are therefore essential. Previous studies have demonstrated the value of qualitative methods such as focus groups to identify clinician priorities and barriers to implementation^4,6,7^. However, most reported systems are evaluated under stable operating conditions, with limited evidence describing how clinician requirements evolve, especially during periods of extreme operational stress.

The COVID-19 pandemic placed unprecedented strain on hospital services and rapidly altered clinical workflows, staffing models, and risk tolerance. In this context, research tools, such as wearable monitoring systems were deployed to help frontline clinical staff, including in our local NHS trust where we deployed our Virtual High Dependency Unit (VHDU) system into clinical practice^8^. This created a unique opportunity to study how clinician perceptions of system features change when technologies are deployed under real-world crisis conditions.

This paper reports findings from a multi-phase, iterative development process for a hospital wearable WMS, combining clinician focus groups conducted before, during, and after the COVID-19 pandemic with quantitative wearability testing. The objectives were to:

1. Identify clinician-perceived priorities for wearable WMS design.
2. Examine how these priorities changed following pandemic deployment.
3. Validate device selection through structured wearability assessment.

### 1.1 Initial system development

Development and testing of the VHDU system was planned in several phases (figure 1). Device accuracy and wearability studies informing development of the vHDU system have been reported previously. As a summary, we sourced all wearable pulse oximeters and chest patches which met pre-specified criteria for consideration in the system. We tested these for wearability on healthy volunteers using a validated wearability questionniare^9^. We tested the accuracy of devices by simulating common patient movements and inducing hypoxia in healthy volunteers^10,11^. We also discussed current monitoring practices and views of wearable monitoring with local clinical staff and patients^4^. We considered the results of these studies, alongside assessments of reliability, connectivity and usability based on observations during these two studies, to select the final devices (one pulse oximeter and one chest patch) for inclusion in the system. We also added a blood pressure cuff to the system (additional testing was not undertaken as only one device was suitable for system integration).

**Figure 1.**
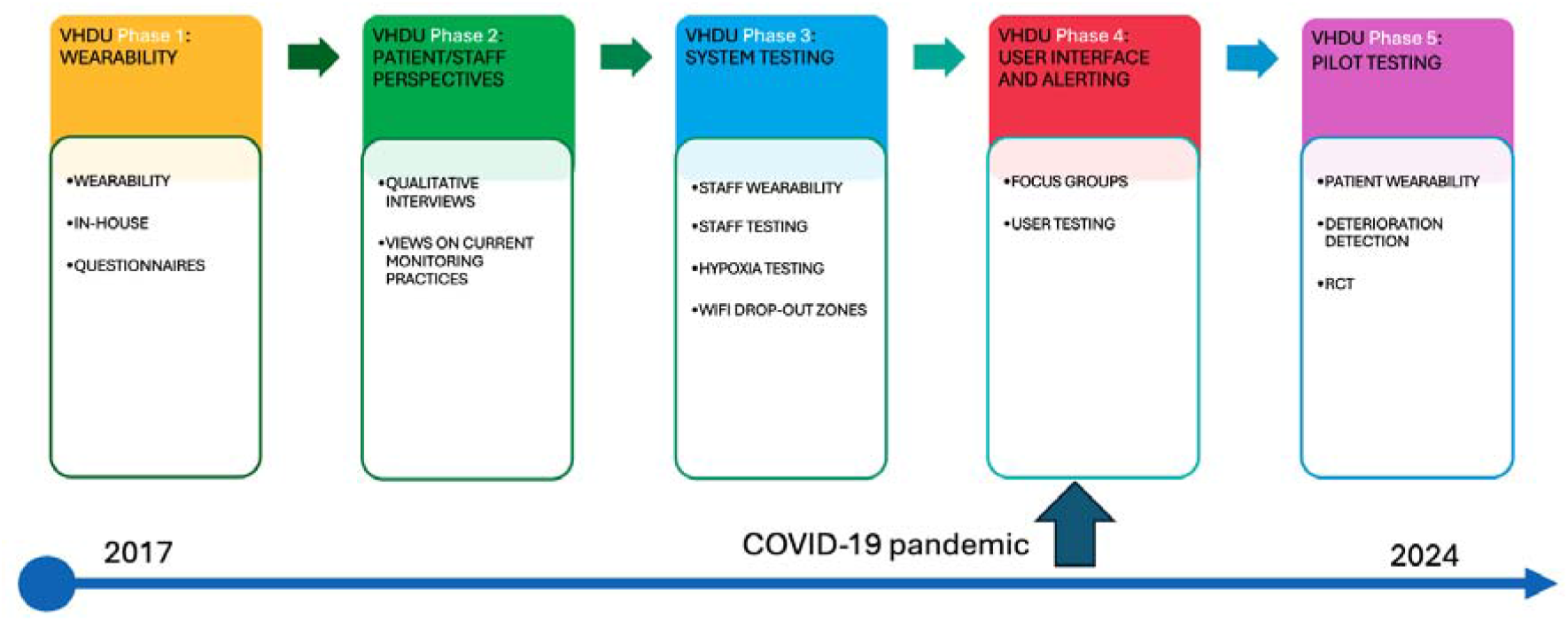
Phases of the virtual High Dependency Unit Wearable Monitoring System

## 2. METHODS

Once the devices had been selected, we co-developed the user interface with clinical staff, to ensure we optimised the display of data generated by the devices for clinical use. The initial prototype user interface was developed by the research team based on existing clinical and engineering knowledge and experiences from previous similar studies^12,13^. We then planned to seek staff feedback on this prototype interface in several phases, in preparation for deployment into clinical practice for the trial.

### 2.2 Initial staff focus groups

Initial focus groups were held with clinical staff on the ward where the RCT would be run. Four members of the research team ran the focus groups: two researchers (one with training in qualitative research methods) and the two engineers developing the system. The research team started by introducing the concept of wearable monitoring systems and demonstrating the devices. The prototype user interface was then presented to clinical staff and a guided discussion followed, based on a topic guide. The focus groups considered the use case for devices, how these could be used in clinical practice, and what should be included in the user interface. The focus group discussions were audio recorded, transcribed verbatim and summarised using content analysis^14^. A report of feedback points was produced and considered by the engineering team in guiding adjustments made to the system.

### 2.3 Pandemic deployment

At this stage in the development of the VHDU system, the COVID-19 pandemic reached the UK. Locally, all research activity not related to the pandemic was paused and the system was deployed on the local infection diseases ward, an isolation ward, to support the pandemic response. The research team worked closely with the ward staff to optimise the system and enable remote monitoring of patients in the isolation ward, and feedback was sought and quickly integrated into the interface over the course of the first two weeks of deployment, with the aim of streamlining useability in a highly pressured environment. We evaluated implementation of the system as a service evaluation and published the views of nursing staff using the system^15^.

### 2.4 Pre-trial refinement focus groups

Following the pandemic, we were able to resume plans to commence the RCT. To ready the system, further focus group sessions were held following the same format as previously. The system was presented to ward clinical staff, including some who had been members of previous focus groups, and the changes made during the pandemic deployment presented and discussed. Further feedback on refinements to the system were sought. Focus groups discussions were again audio recorded, transcribed and analysed using content analysis to summarise the main points for consideration.

### 2.5 In-trial wearability testing

During the RCT, device patient wearability was assessed using the same validated wearability questionnaire as used during initial wearability testing of candidate devices in healthy volunteers (VHDU phase 1)^9^. Participants were asked to complete the questionnaire either on paper or with a member of the research delivery team after the devices had been removed. The Comfort Rating Scale (CRS)^16^ includes statements related to six domains (emotions, attachment, harm, perceived change, movement and anxiety), with some domains including more than on statement. Participants are asked to rate each statement on a Likert scale from 0 (strongly disagree) to 20 (strongly agree). Most questions were negatively framed, with a lower score indicating a more positive view of the wearability of the devices. However, statement 3 in the emotions domain is framed positively therefore a higher score indicates a more positive view of wearability. Data were recorded in RedCap as part of the trial Case Report Form and analysed descriptively.

### 2.6 Ethical approval

This manuscript reports findings from multiple phases of the vHDU project related to system development, with separate approvals secured for each phase. For the user interface development (phase 4) South Central Hampshire A Research Ethics Committee granted ethical approval (reference 19/SC/0181) and we registered the protocol (SRCTN51530527). For the pandemic service evaluation work ethical approval was not required, but the project was peer reviewed locally and registered with the quality improvement team (reference: Datix 5973). For the in-trial wearability assessment within the pilot RCT (phase 5) Wales Research Ethics Committee 5 granted ethical approval (ref: 21/WA/0250). Participants in phase 4 and phase 5 provided written informed consent prior to participating in the focus groups and wearability questionnaire study respectively.

## 3. RESULTS

### 3.1 Initial focus groups (pre-pandemic)

Three focus group sessions were held on 14^th^ November 2019 with a total of 15 members of staff: 8 nurses, 1 clinical support worker (nursing assistant) and 6 allied health professionals, with varying durations of clinical experience. The engineers used this initial feedback to make adjustments to the system.

After deployment of initial adjustments based on feedback, a second round of three focus group discussions was held on 12^th^ March 2020 with 11 members of staff: 10 nurses and 1 clinical support worker (some of whom had attended a previous focus group session), following the same format. The revised user interface was presented to clinical staff, and specific feedback sought by the research team, focusing on how the interface would be used in practice. General feedback was also sought. Key points from both focus group rounds are presented in table 1 and were considered in the development of the system.

**Table 1.** Discussion points and feedback from initial user interface development focus groups.

| Discussion point | Summary of feedback |
| --- | --- |
| Audible alerts | Only at nurses station, not at bedside<br>Need for flexibility for night versus day sound levels<br>Concern about alarm fatigue |
| Position of alerts | Strongly in favour of main screen at nurses station<br>Addition of either phone-type device in pocket vibrating or tablet at bedside/outside bay flashing<br>Wall-mounted tablets favoured over bedside tables<br>Concerns raised around patient confidentiality<br>Strong negative feedback to accessing alerts on computers on wheels or other desktop devices |
| Idea of phone-type device in pocket explored further | Initially overall positive<br>Co-ordinator should have overview of all alerts on ward<br>Concerns about perceptions of professionalism if frequently looking at a phone-type device<br>In second round of focus groups concept of phone-type device was explored in more detail and consensus was to reject as practicalities considered and multiple barriers identified |
| Dwell-time of intermittent measurements (blood pressure and temperature) | Need clear highlighting that intermittent reading, not current data<br>Clear discomfort regarding any dwell time for intermittent vital signs<br>Perception that availability of respiratory rate values from chest patch will improve time efficiency of vital-sign measurements |
| Flexibility of parameters | Need to alter alerts for e.g. COPD patients (accounted for in NEWS 2) |
|  | Flexibility of parameters to avoid alert fatigue?<br>Suggested facility to annotate and pause alerts e.g. during physiotherapy where vital signs may change temporarily? |

Initial discussions focused on audible alerts, the potential for a nurse-held personal device to received data and alerts, and dwell-time for intermittently measured vital signs (not provided by the VHDU system).

### 3.2 Deployment and feedback during the COVID-19 pandemic

Table 2 outlines feedback from clinical staff using the system to manage patients hospitalised with COVID-19. Changes were focused on improving usability so that clinical staff could become familiar with, and use, the system with minimal training and support. This included removing the manual calibration period for the chest patch, adding clear labelling and a dedicated screen at the nurses station for viewing vital sign data.

**Table 2.** Staff feedback from deployment during the COVID-19 pandemic and actions taken.

| Feedback | Changes made to system in response |
| --- | --- |
| Need to prevent patients tampering with tablet | Labels were made for the tablets and plugs |
| The window displaying the central station on the main screen at the nursing station often closed by clinical staff using other applications | Dedicated screen installed to display the central screen of the system only |
| Staff raised concern that a patient would not be able to walk 35 steps to calibrate the chest-patch device | Adjustment to calibration process with no need for patient to walk |
| Concerns raised regarding the pulse oximeter under-reading | Team training repeated. Team in agreement that under-reading is safer than over-reading |
| Concerns about inaccuracy of the skin temperature reading from the patch (previous version of chest patch than the one used for the RCT which displayed body temperature) | Skin temperature removed from display as misleading |
| Nurses do not seem to be entering partial observations into the electronic patient record | Possibly due to downgrading of PPE, hence nurses now more willing to enter room and take observation sets |
| Staff would like audible alerts added to the system | For consideration in future iterations of the system |
| Did not want a personal device (e.g. phone or personal computer) to review system and alerts | Did not pursue personal phone option further |

### 3.3 Refinement for deployment onto the surgical ward – post-pandemic

Three further focus group sessions were held with clinical staff on the trial ward in April and May 2021, with eight attendees (seven ward nurses with varying experience and one clinical support worker). The refined interface was shared with clinicians, along with research staff perspectives of how the system was used in practice. Table 3 presents a summary of feedback areas and responses. All suggestions were considered and implemented into the system where possible. It was not possible to add a facility to save an ECG trace for later reference, although it is now possible to scroll back through the waveforms. To ensure patient safety, the facility to adjust the alert parameters was not deployed, but alerts were instead based on the local EWS system. In response to the lack of consensus on duration of silencing alerts, three options were offered (15 minutes, 30 minutes and 60 minutes), with 60 minutes the maximum time an alert could be silenced. The final system is shown in figures 2, 3 and 4.

**Table 3.**
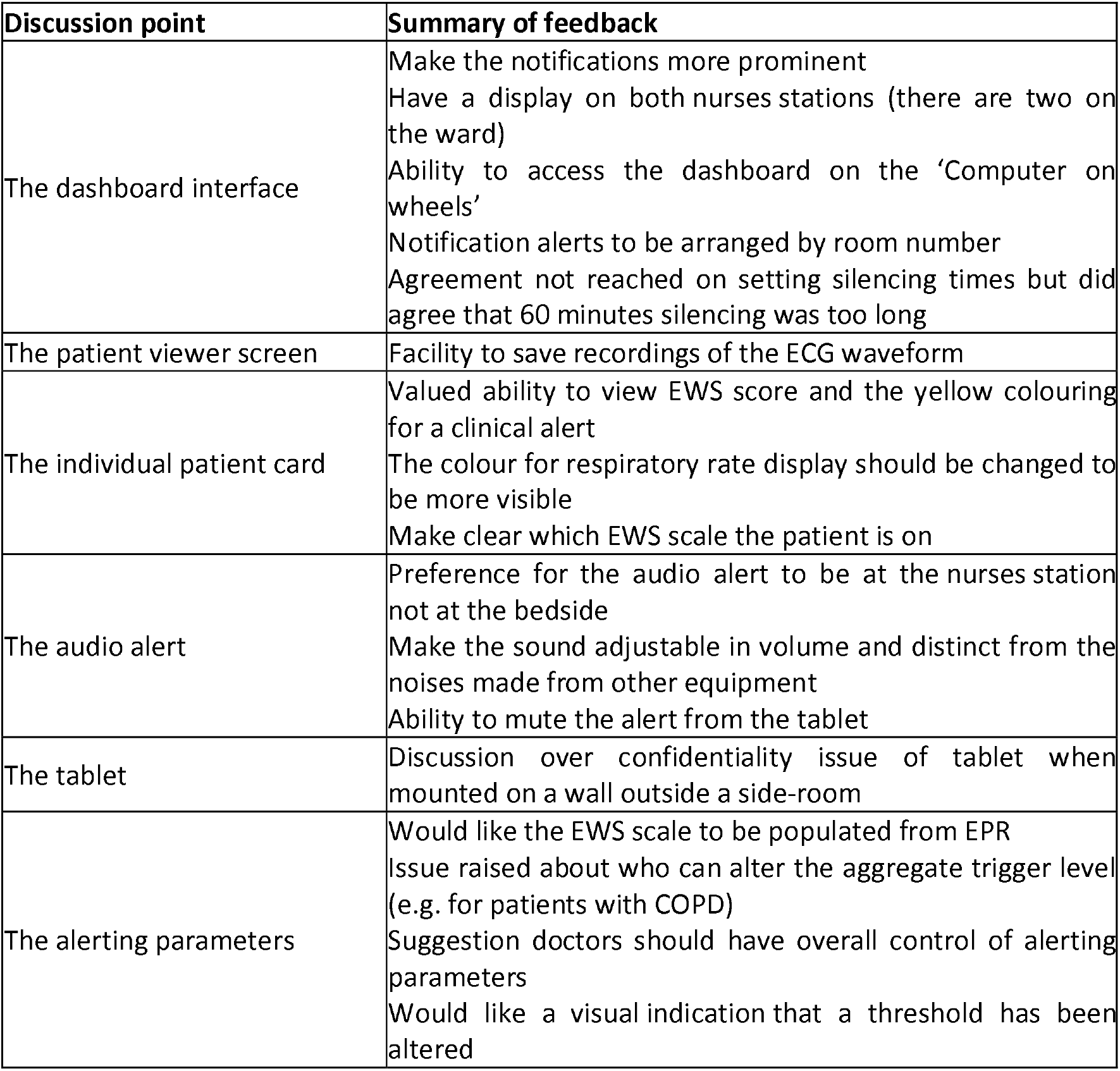
Summary of feedback from second round of focus groups in preparation for trial deployment.

**Figure 2.**
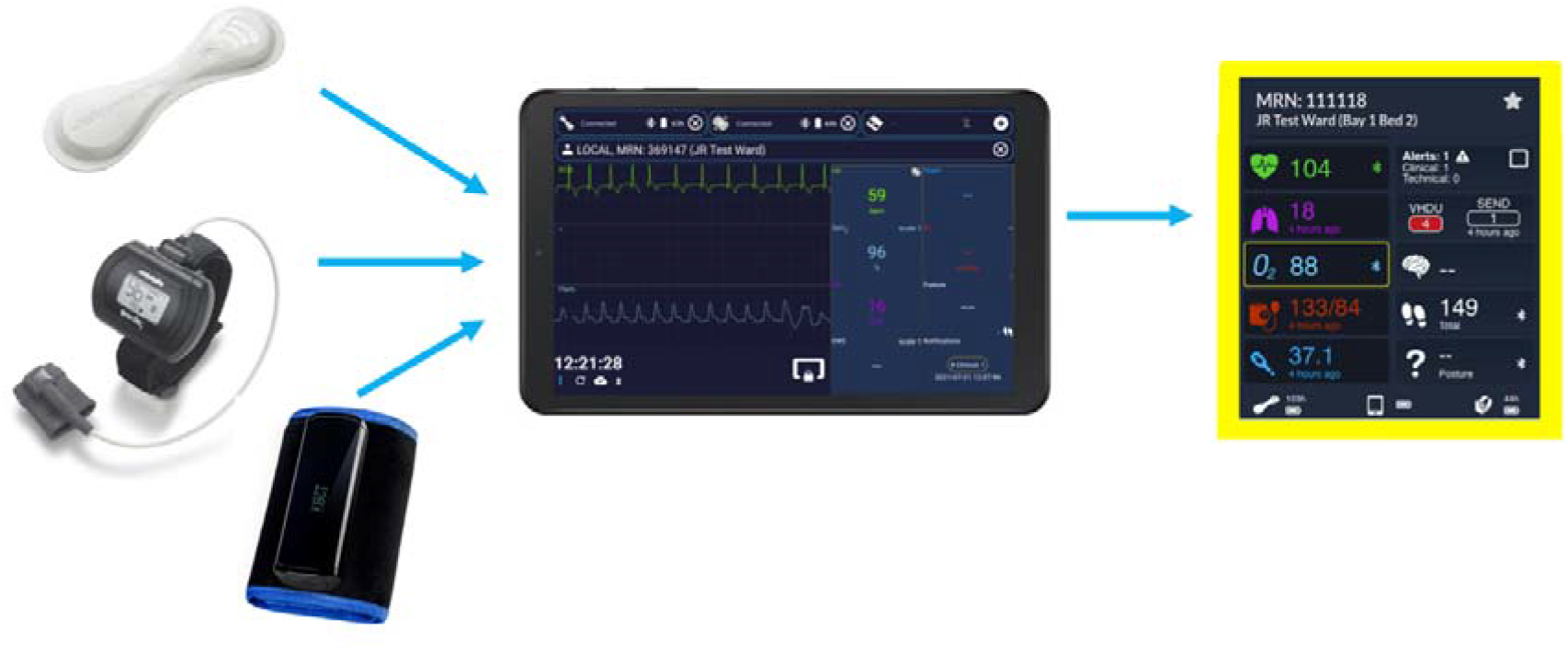
Overview of the vHDU WMS components with the patient card alerting at the nurse dashboard.

**Figure 3.**
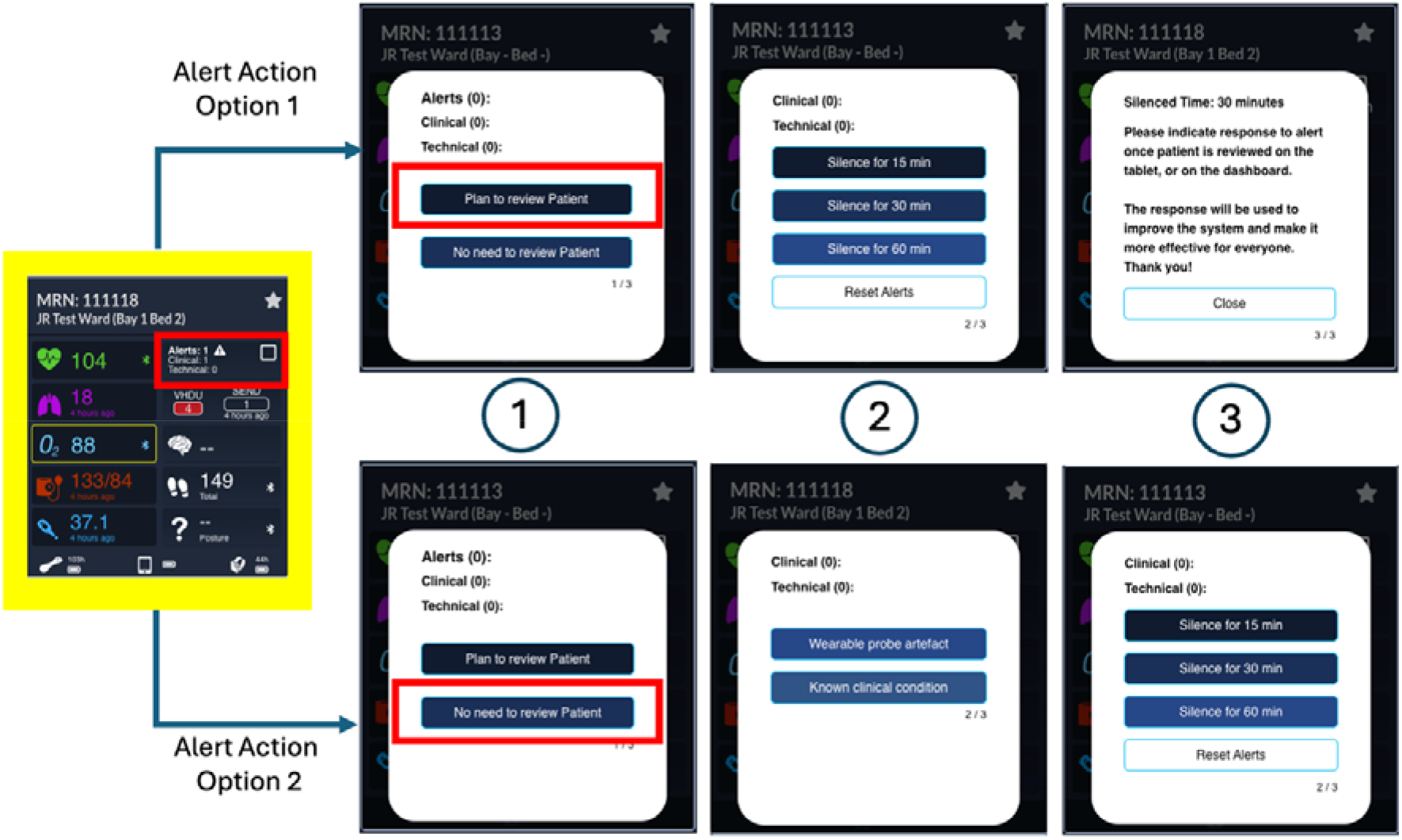
Collage of various images of the vHDU Alert system (main dashboard and alerting scenario)

**Figure 4.**
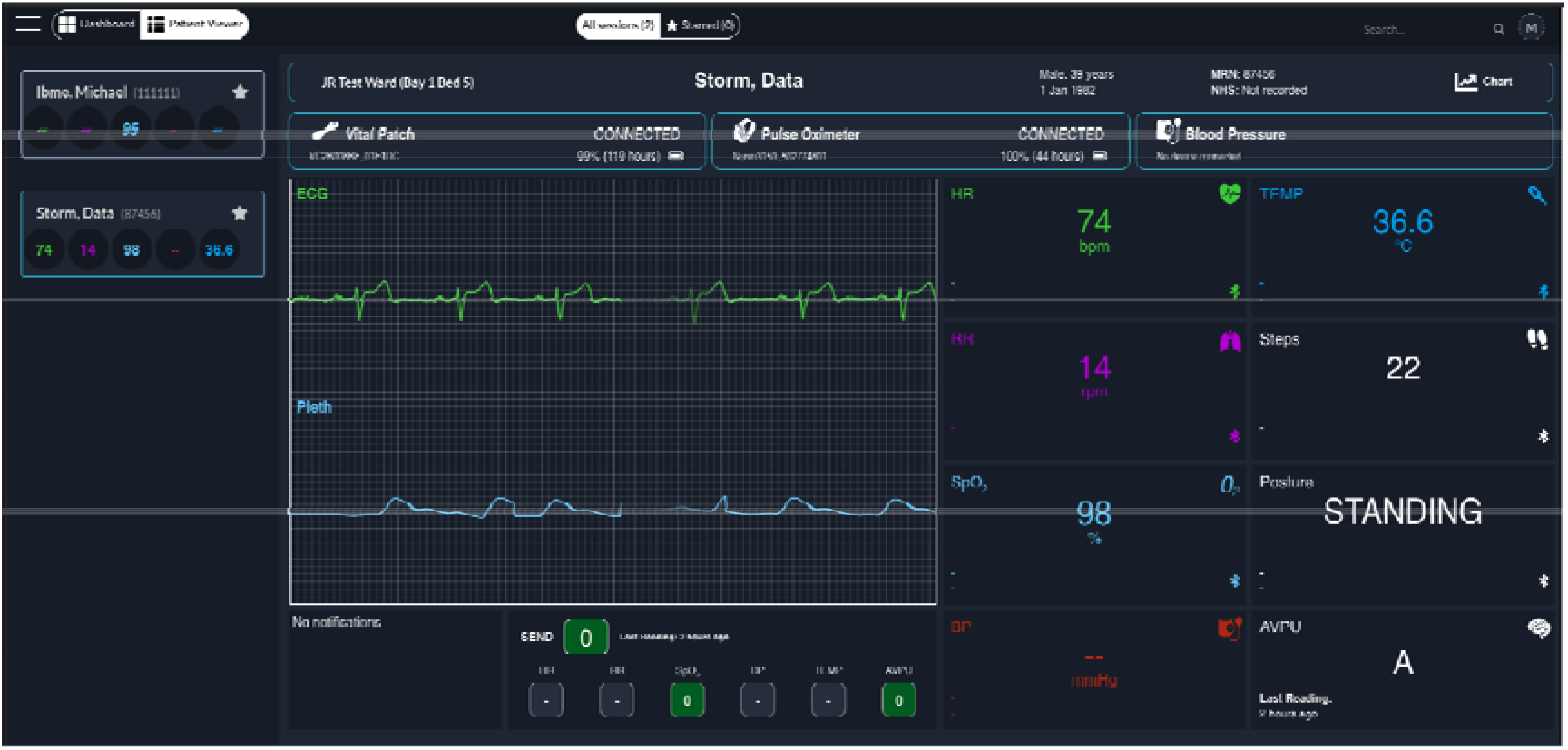
vHDU Clinician Patient Viewer (showcasing live ECG and PPG)

### *3*.4 Wearability Testing

A total of 169 out of 235 participants completed the wearability questionnaire. Only 21 completed the questions for the blood pressure cuff as this was quickly removed due to consistently reported discomfort from the first 31 patients. Data on comfort rating scores are presented in table 4 below, with median scores displayed as Strongly disagree=1-5 (dark green); disagree=6-9 (green); neither=10-13 (grey); agree=14-17 (red); and strongly agree=18-21 (dark red). The blood pressure cuff was poorly tolerated (with median scores for 6 out of 14 statements either agree or strongly agree). Almost all statements for the other two devices were strongly disagreed with, indicating good wearability (table 4).

**Table 4.**
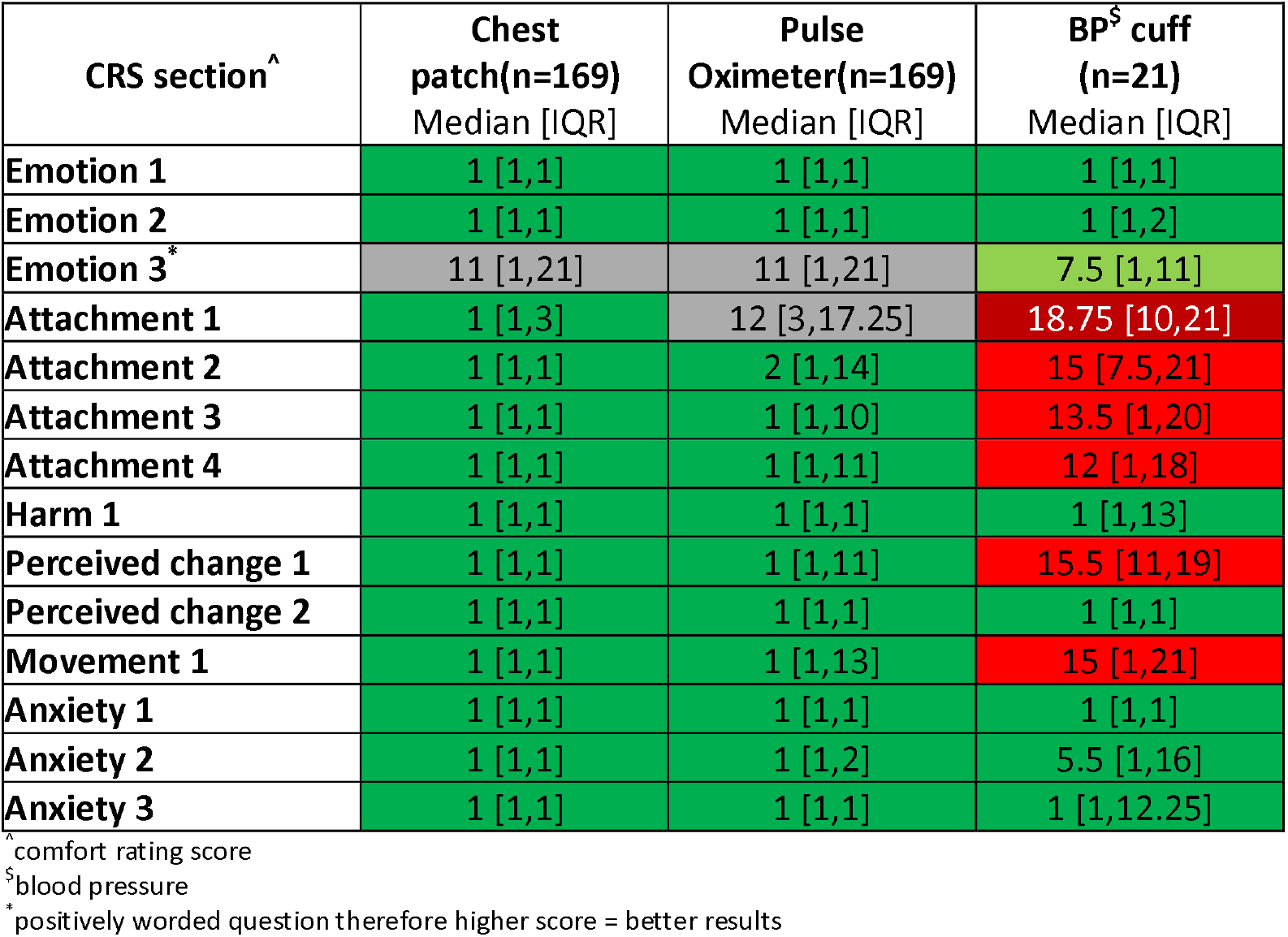
Comfort rating scores for each device.

| CRS section<sup>^</sup> | Chest patch(n=169)<br>Median [IQR] | Pulse Oximeter(n=169)<br>Median [IQR] | BP<sup>§</sup> cuff (n=21)<br>Median [IQR] |
| --- | --- | --- | --- |
| Emotion 1 | 1 [1,1] | 1 [1,1] | 1 [1,1] |
| Emotion 2 | 1 [1,1] | 1 [1,1] | 1 [1,2] |
| Emotion 3<sup>*</sup> | 11 [1,21] | 11 [1,21] | 7.5 [1,11] |
| Attachment 1 | 1 [1,3] | 12 [3,17.25] | 18.75 [10,21] |
| Attachment 2 | 1 [1,1] | 2 [1,14] | 15 [7.5,21] |
| Attachment 3 | 1 [1,1] | 1 [1,10] | 13.5 [1,20] |
| Attachment 4 | 1 [1,1] | 1 [1,11] | 12 [1,18] |
| Harm 1 | 1 [1,1] | 1 [1,1] | 1 [1,13] |
| Perceived change 1 | 1 [1,1] | 1 [1,11] | 15.5 [11,19] |
| Perceived change 2 | 1 [1,1] | 1 [1,1] | 1 [1,1] |
| Movement 1 | 1 [1,1] | 1 [1,13] | 15 [1,21] |
| Anxiety 1 | 1 [1,1] | 1 [1,1] | 1 [1,1] |
| Anxiety 2 | 1 [1,1] | 1 [1,2] | 5.5 [1,16] |
| Anxiety 3 | 1 [1,1] | 1 [1,1] | 1 [1,12.25] |

<sup>^</sup> comfort rating score
<sup>§</sup> blood pressure
<sup>\*</sup> positively worded question therefore higher score = better results

## 4. DISCUSSION

This study demonstrates the value of iterative, clinician-led development to maximise usability of wearable monitoring systems in clinical practice, and provides insight into how extreme clinical contexts can reshape perceived system requirements. The addition of a patient wearability assessment study marks the completion of our system testing, ensuring accuracy and patient acceptability as well as usability. This careful preparatory work resulted in a system which was acceptable to both patients and staff^15^, and was successfully used in a 240 patient pilot RCT.

### Evolution of Interface and Alerting Preferences

Early focus group sessions, conducted prior to the COVID-19 pandemic, suggested strong interest in flexible access to alerts, including exploration of smartphone-type devices carried by clinicians. While initially viewed as a potential solution to improve responsiveness, concerns regarding professionalism, distraction, infection control, and cognitive load emerged even at this stage. Following research team learning during real-world deployment during the pandemic, clinician preferences shifted away from personal or mobile devices. Fixed, ward-based displays and audible alerts located at the nurses’ station were preferred, reflecting the realities of high workload, PPE use, and the need for shared situational awareness. This shift underscores that perceived usefulness of technologies cannot be reliably assessed without exposure to authentic clinical pressures. While pre-pandemic focus groups expressed strong negative feedback towards accessing alerts on computers on wheels or desktop devices, post-pandemic feedback prioritised audibility and central visibility over device type. This suggests that during crisis conditions, immediacy and shared awareness outweigh concerns about interface modality.

### Alarm Fatigue and Parameter Flexibility

Across all phases of the testing, clinicians consistently emphasised the risk of alarm fatigue, also identified in previous research^1,17–19^. Requests for adjustable alert thresholds, silencing options, and contextual annotation reflected a desire for systems that support clinical judgement rather than enforce rigid automation. However, maintaining patient safety required careful limitation of user-modifiable parameters. The final design, which aligned alerting behaviour with the local Early Warning Score system, represents a compromise between flexibility and governance.

### Wearability as a Prerequisite for Adoption

Patient wearability testing confirmed our previous findings in healthy volunteers^9^ that the chest patch and pulse oximeter were well tolerated over extended periods, supporting their suitability for continuous monitoring. In contrast, the blood pressure cuff demonstrated poor tolerability across multiple domains, leading to its removal from the system. This highlights the importance of evaluating not only sensor accuracy but also physical burden when designing multi-device monitoring solutions.

### Implications for Engineering Design

From an engineering perspective, these findings emphasise that WMS design is not a static optimisation problem. User requirements evolve in response to clinical context, workload, and external stressors. Systems intended for routine care must be resilient to rapid repurposing during emergencies, and design decisions should prioritise adaptability, simplicity, and shared visibility over feature proliferation.

## 5. CONCLUSION

This work describes the iterative development of a hospital wearable remote monitoring system informed by clinician focus groups and wearability testing across pre-pandemic, pandemic, and post-pandemic phases. Clinician priorities consistently centred on alert visibility, alarm fatigue mitigation, and workflow integration, while preferences for access mechanisms and alert modalities changed substantially following real-world deployment during COVID-19. Our results demonstrate that clinician perceptions of “ideal” system features can differ markedly from those formed under routine conditions once technologies are exposed to operational stress. Incorporating longitudinal qualitative feedback alongside quantitative wearability assessment enabled the development of a system that was both clinically acceptable and technically robust.

These findings support the use of iterative, user-centred design methodologies in the engineering of safety critical healthcare technologies and highlight the importance of evaluating systems under realistic clinical conditions prior to large scale trials.

## Data Availability

All data produced in the present work are contained in the manuscript

## AKNOWLEDGEMENTS

We would like to acknowledge the research team who supported this work throughout the different phases of development, including Peter Watkinson, Mauro Santos, Marco Pimentel, Carlos Areia, Chris Biggs, Holly Edmundson, Louise Young and Annika Jarman. We also gratefully acknowledge the patients who participated in the RCT and completed wearability questionnaires, and the clinical staff who willingly and rapidly adopted the system into their practice under extremely difficult circumstances during the COVID-19 pandemic.

## REFERENCES

1. Areia, C. et al. The impact of wearable continuous vital sign monitoring on deterioration detection and clinical outcomes in hospitalised patients: a systematic review and meta-analysis. Crit. Care 25, 1–17 (2021).

2. Khanna, A. K., Flick, M. & Saugel, B. Continuous vital sign monitoring of patients recovering from surgery on general wards: a narrative review. Br. J. Anaesth. 134, 501–509 (2025).

3. Downey, C. L., Chapman, S., Randell, R., Brown, J. M. & Jayne, D. G. The impact of continuous versus intermittent vital signs monitoring in hospitals: A systematic review and narrative synthesis. Int. J. Nurs. Stud. 84, (2018).

4. Areia, C. et al. Experiences of current vital signs monitoring practices and views of wearable monitoring: A qualitative study in patients and nurses. J. Adv. Nurs. 00, 1–13 (2021).

5. Downey, C. L., Brown, J. M., Jayne, D. G. & Randell, R. Patient attitudes towards remote continuous vital signs monitoring on general surgery wards: An interview study. Int. J. Med. Inf. 114, 52–56 (2018).

6. Kooij, L., Peters, G. M., Doggen, C. J. M. & van Harten, W. H. Remote continuous monitoring with wireless wearable sensors in clinical practice, nurses perspectives on factors affecting implementation: a qualitative study. BMC Nurs. 21, 53 (2022).

7. van Noort, H. H. J. et al. Three Years of Continuous Vital Signs Monitoring on the General Surgical Ward: Is It Sustainable? A Qualitative Study. J. Clin. Med. 13, 439 (2024).

8. Santos, M. D. et al. A Real-Time Wearable System for Monitoring Vital Signs of COVID-19 Patients in a Hospital Setting. Front. Digit. Health 0, 120 (2021).

9. Areia, C. et al. Wearability testing of ambulatory vital sign monitoring devices: Prospective observational cohort study. JMIR MHealth UHealth 8, (2020).

10. Areia, C. M. et al. A Chest Patch for Continuous Vital Sign Monitoring: Clinical Validation Study During Movement and Controlled Hypoxia. J Med Internet Res *2021239e27547 Httpswwwjmirorg20219e27547* 23, e27547 (2021).

11. Vollam, S. et al. Wearable pulse oximeters in the prompt detection of hypoxaemia and during movement: a diagnostic accuracy study. J. Med. Internet Res. (2021).

12. Watkinson, P. J. et al. A randomised controlled trial of the effect of continuous electronic physiological monitoring on the adverse event rate in high risk medical and surgical patients. Anaesthesia 61, 1031–1039 (2006).

13. Watkinson, P. J. et al. Early detection of physiological deterioration in post-surgical patients using wearable technology combined with an integrated monitoring system: a pre- and post-interventional study. 2020.12.01.20240770 Preprint at 10.1101/2020.12.01.20240770 (2020).

14. Hsieh, H.-F. & Shannon, S. E. Three Approaches to Qualitative Content Analysis. Qual. Health Res. 15, 1277–1288 (2005).

15. Buss, A. et al. Using a novel ambulatory monitoring system to support patient safety on an acute infectious disease ward during an unfolding pandemic. J. Adv. Nurs. 80, 2452–2461 (2024).

16. Knight, J. F. & Baber, C. A Tool to Assess the Comfort of Wearable Computers. Hum. Factors 47, 77–91 (2005).

17. Weller, R. S., Foard, K. L. & Harwood, T. N. Evaluation of a wireless, portable, wearable multi-parameter vital signs monitor in hospitalized neurological and neurosurgical patients. J. Clin. Monit. Comput. 32, 945–951 (2018).

18. Downey, C., Randell, R., Brown, J. & Jayne, D. G. Continuous versus intermittent vital signs monitoring using a wearable, wireless patch in patients admitted to surgical wards: Pilot cluster randomized controlled trial. J. Med. Internet Res. 20, e10802 (2018).

19. Edwards, C. et al. “You can’t be with your patients all the time”: Patient and staff views of a wearable vital signs monitoring system. J. Adv. Nurs. online ahead of print, (2026).

